# Ryanodine Receptor Inhibition with Dantrolene Prevents Ventricular Tachycardia Induction in Patients with Structural Heart Disease – A Randomized Controlled Trial

**DOI:** 10.1101/2025.08.17.25333868

**Authors:** Majd A. El-Harasis, Zachary T. Yoneda, Bibin Varghese, Dakota Grauherr, Diane M. Crawford, Jeffrey Schmeckpeper, Hollie L. Williams, Fei Ye, Lili Sun, Harikrishna Tandri, Travis Richardson, Arvindh Kanagasundram, Giovanni Davogustto, Dan Roden, Michael Mantinan, Mias Pretorius, Frederic T. Billings, Kara Siegrist, Wendell S. Akers, William G. Stevenson, Bjorn C. Knollmann, M. Benjamin Shoemaker

**Author notes:** **Correspondence**: M. Benjamin Shoemaker, MD, MSCI, Department of Medicine, Division of Cardiovascular Medicine, Vanderbilt University Medical Center, 1211 Medical Center Drive Nashville, TN 37232, U.S.A., Bjorn C. Knollmann, M.D., Ph.D., Department of Medicine, Division of Genetic Medicine and Clinical Pharmacology, Vanderbilt University Medical Center 1211 Medical Center Drive Nashville, TN 37232, U.S.A. **Clinical Trial Registration:** <u>Study Details | A Clinical Trial Utilizing Dantrolene in Patients with Ventricular Arrythmias. | ClinicalTrials.gov</u> (NCT04134845). Date of Registration: October 18, 2019.

## Abstract

Despite clinical need, no new drugs for treating ventricular tachycardia (VT) have emerged for over 20 years. In structural heart disease, posttranslational modifications render intracellular ryanodine receptor-2 (RyR2) calcium release channels leaky, which increases VT risk. Treatment with dantrolene, an RyR2 inhibitor, prevents VT in animal models. Here, we test Dantrolene in a randomized, double-blinded, placebo-controlled clinical trial of 51 patients with structural heart disease undergoing catheter ablation for ventricular arrhythmias. Cardiac electrophysiologic parameters, hemodynamics and VT inducibility were assessed at baseline and after IV administration of dantrolene (29 patients) or placebo (22 patients). Approximately half of study participants were inducible at baseline. Dantrolene reduced inducible VT by 66% whereas placebo had no effect (odds ratio 0.23, 95% CI 0.06-0.90, P=0.034). Dantrolene had no significant effects on conduction velocity, effective refractory periods, heart rate, ECG intervals, blood pressure, or cardiac function. We conclude that Dantrolene reduced VT inducibility in high-risk patients with a favorable safety profile. These findings support the safety and efficacy of targeting RyR2 for arrhythmia prevention in humans. *Clinicaltrials.gov NCT04134845*.

## INTRODUCTION

Sudden cardiac death (SCD) accounts for up to 20% of deaths world-wide, with ventricular tachycardia (VT) and ventricular fibrillation (VF) from structural heart disease (e.g., ischemic and non-ischemic cardiomyopathy) as major causes.^1,2^ VT also increases the risk of heart failure hospitalizations, and once VT occurs, it is likely to recur.^3–5^ Antiarrhythmic drugs (AADs) have been used for decades to treat VT/VF, but their clinical utility is limited by toxicity and poor efficacy, especially in patients with structural heart disease.^6^ No new AADs have become available in over 20 years.^7^ Hence, a major unmet need is the development of a new class of AADs to prevent VT/VF in structural heart disease.

Intracellular calcium leak via the cardiac type 2 ryanodine receptor (RyR2) has been implicated as a mechanism contributing to VT/VF risk in patients with genetic arrhythmia syndromes as well as in patients with structural heart disease, making RyR2 inhibition a promising antiarrhythmic drug target.^8,9^ RyR2 block by class IC AADs flecainide and propafenone is clinically effective for VT prevention in patients carrying disease mutations that render RyR2-calcium release channels leaky.^10,11,12^ However, in structural heart disease, the prominent sodium channel blockade of class IC drugs slows conduction velocity, which can promote reentrant VT and increase mortality.^6^ The only other RyR2 inhibitor in clinical use is dantrolene, which is currently approved to treat malignant hyperthermia acutely and muscle spasticity chronically. Dantrolene is a non-selective RyR inhibitor that blocks the type 1 RyR receptor present in skeletal muscle (RyR1) and the RyR2 receptor present in cardiac muscle.^13^ Dantrolene reduces susceptibility to VT/VF in experimental models of VF, heart failure, and myocardial infarction.^14,15^ Dantrolene appears to reduce VT/VF incidence in an uncontrolled study of patients at risk for VT/VF storm,^16^ but it has not yet been studied in a controlled trial. Here, we report a randomized placebo-controlled trial that investigates the efficacy, safety, and electrophysiologic effects of RyR2 inhibition with dantrolene in patients with structural heart disease and high risk for VT.

## RESULTS

### Efficacy endpoints

From December 2020 to March 2024, a total of 68 participants were randomized to dantrolene (N=41) or placebo (N=27, **Supplemental Figure 4**). Seventeen were withdrawn after randomization for safety concerns related to the clinical ablation procedure (i.e., concerns regarding hemodynamic stability) prior to commencement of the research protocol. Study withdrawals occurred without knowledge of the randomization results. All 51 participants that entered the research protocol also completed the research protocol (29 in the dantrolene group and 22 in the placebo group). **Table 1** lists the demographics and baseline clinical characteristics. Differences between treatment assignments were statistically compared. Using a Bonferroni corrected P=value of <0.002 (0.05/27), there were no statistically significant differences between groups.

**Table 1:**
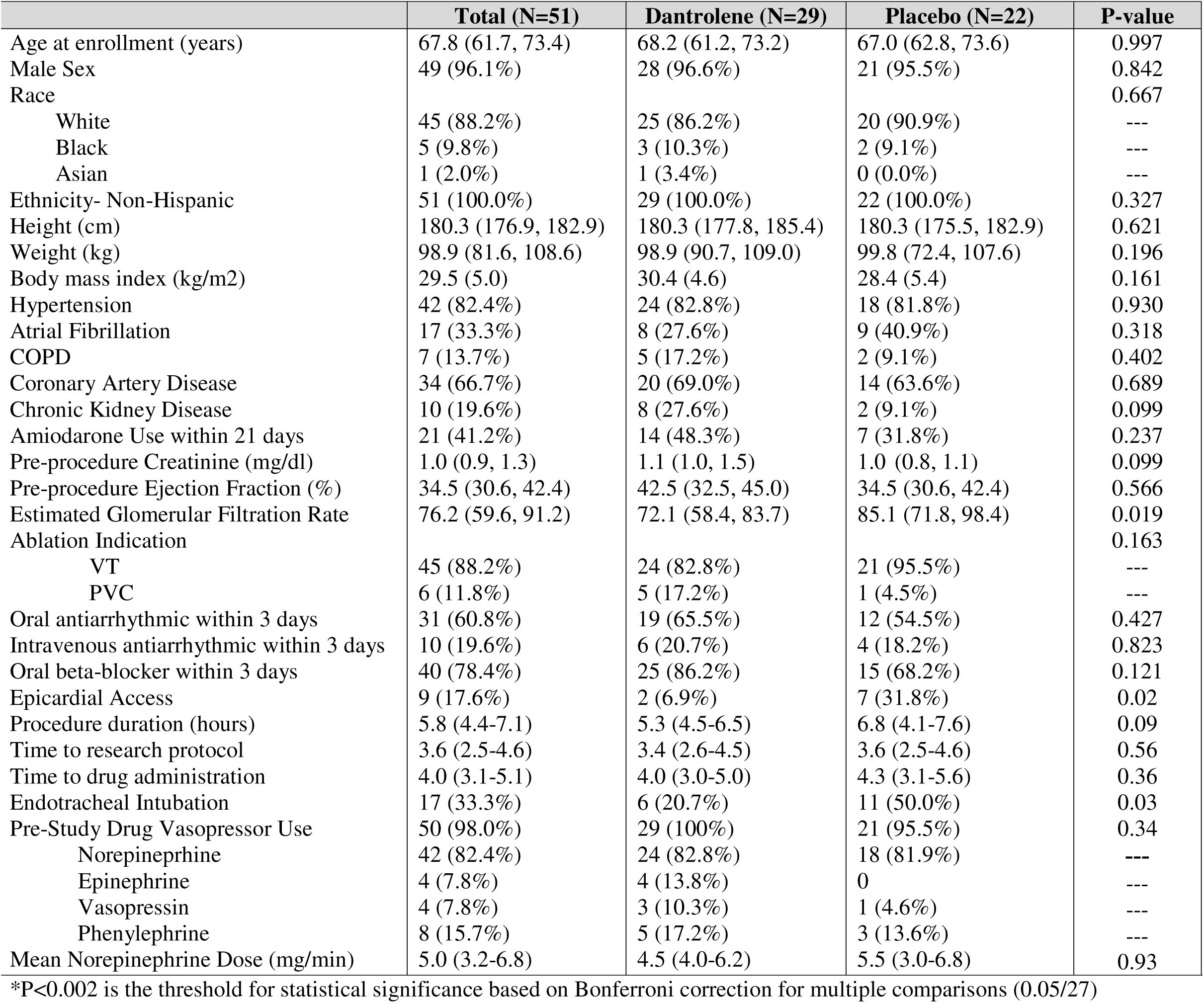
Baseline characteristics and ablation details of participants who completed the protocol.

Before study drug, 22 participants (43.1%) had inducible VT (**Supplemental Table 3**). In the dantrolene group, 12 participants (41.4%) were inducible pre-drug administration compared to 4 participants (13.8%) post-drug administration (Δ −27.6%, P=0.04, **Figure 1A**). In the placebo group, 10 participants (45.5%) were inducible pre-drug administration compared to 9 participants (40.9%) post-drug administration (Δ −4.6%, P=1). Compared to placebo, dantrolene reduced inducible VT by 77% (odds ratio 0.23, 95% CI 0.06-0.90, P=0.034 by a logistic regression model, **Figure 1B**). Results from the ordinal inducibility endpoint are presented in **Supplemental Table 3.**

**Figure 1:**
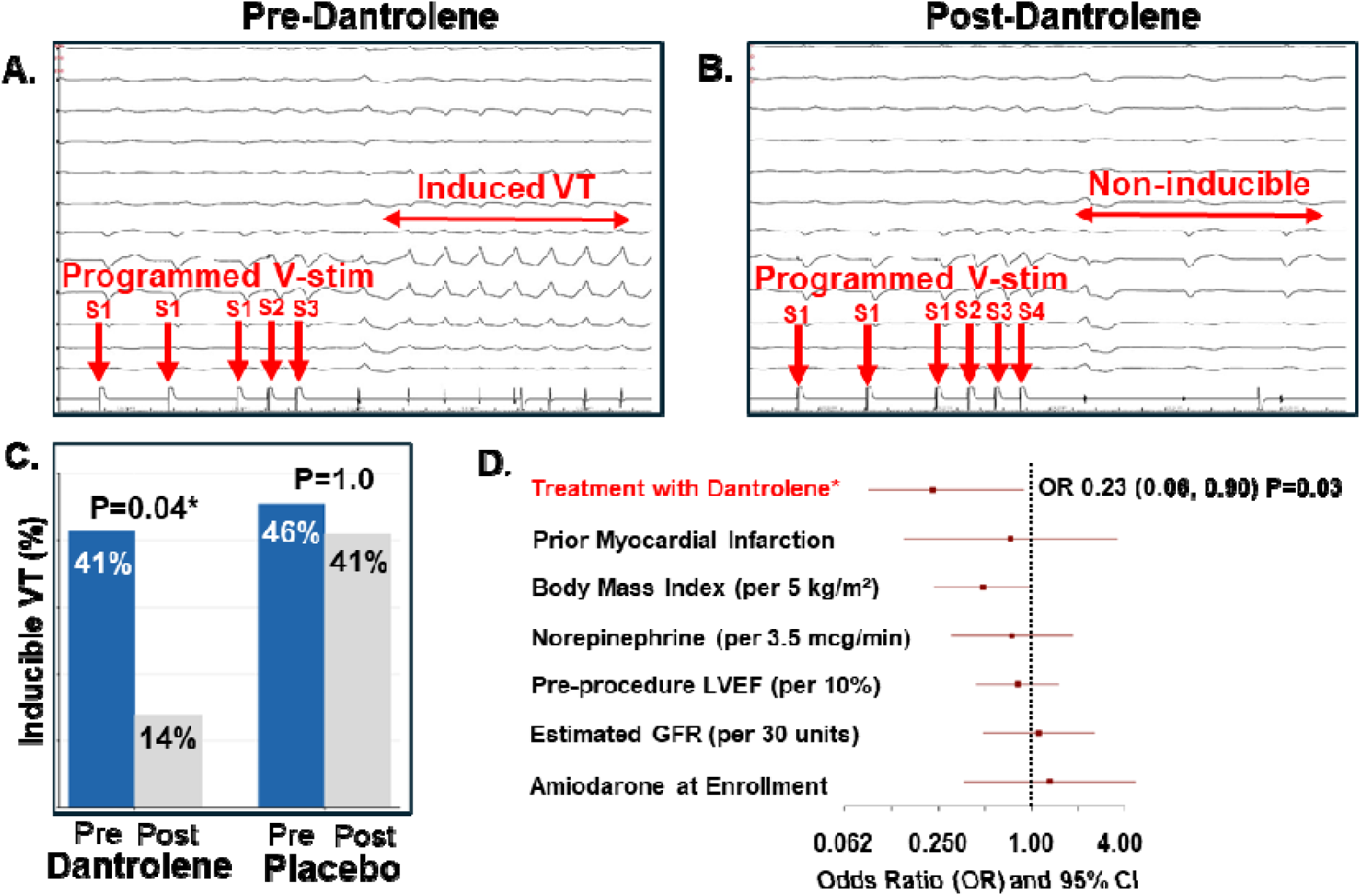
Panels A and B. **Surface electrograms of a participant who was randomized to treatment with dantrolene** and was inducible for sustained monomorphic VT with programmed ventricular-stimulation (V-stim) pre-dantrolene (A) and non-inducible post-dantrolene (B). **C. Bar graph of VT inducibility.** In the dantrolene group, 12 participants (41%) were inducible pre-drug compared to 4 (14%) post-drug (P=0.04). In the placebo group, 10 participants (46%) were inducible pre-drug compared to 9 (41%) post-drug (P=1.0). **D. Univariable analysis of VT inducibility.** Treatment with dantrolene reduced VT inducibility by 77% (OR 0.23 [0.06, 0.90] P=0.03) compared to placebo. Higher BMI was also marginally associated with lower VT inducibility (OR 0.49 [95% C : 0.24, 1.00], P=0.05), but not the other variables listed.

### Safety endpoints

#### Hemodynamics

Dantrolene had no significant effect on blood pressure, cardiac index, pulmonary artery pressure or right atrial pressure (**Supplemental Figures 5, 6**). Mean arterial pressure was analyzed using a linear mixed-effect model (beta −1.3 [95% CI −6.1, 3.6], p=0.6, **Supplemental Figure 7)**. There was no association between dantrolene and cardiac index as analyzed using a linear mixed-effect model (beta=0 [95% CI –0.4, 0.3], p=0.97, **Supplemental Figure 8**). There was no association between dantrolene and post-drug RA pressure (beta 0.54 [−0.64 to 1.72], p=0.37) using a linear regression model with adjustment for pre-drug RA pressure. There was no association between dantrolene and post-drug PA systolic pressure when corrected for multiple comparisons (beta 3.53 [0.19-6.88], p=0.04) using a linear regression model with adjustment for pre-drug PA systolic pressure. There was no association between dantrolene and post-drug PA diastolic pressure (beta 0.99 [-0.9 to 2.9], p=0.31) using a linear regression model with adjustment for pre-drug PA diastolic pressure.

#### Heart Rate and ECG

Heart rate and ECG intervals were measured every two minutes for 20 minutes following drug administration. A mixed-effect model analyzed the association between dantrolene and RR interval (beta 1.8 ± 2.9 [95% CI −3.8, 7.5], p=0.53), PR interval (beta 2.8 [-30.4, 35.9], p=0.87), QRS duration (beta 0.37 [-19.7, 20.4], p=0.97), and QTc interval (beta −19.2 [-40.9, 2.6], p=0.09) and found no significant associations (**Supplemental Figure 9-12)**. Pooled analyses for QRS duration, and QTc interval are displayed in **Supplemental Figures 13, 14**.

#### Conduction and Refractoriness

Pre-study drug, VERP was 280 (260-300) ms in the higher voltage region compared to 285 (255-315) ms in the low voltage region (p=0.03). Baseline conduction time was 54.4 (43.9-74.7) ms in the higher voltage region compared to 76.0 (52.0-98.0) ms in the low voltage region (p=0.01). Conduction restitution curve inflection point was 352.5 (320-371) ms in the higher voltage region compared to 330 (296-362) ms in the low voltage region (p=0.61). There was no significant change in ventricular ERP, baseline conduction time, and conduction restitution curve inflection point in both the higher and low voltage regions pre- vs. post-dantrolene. Pre- and post-drug differences in these values are listed in **Supplemental Table 4**.

#### Other Adverse Events

Five participants (13.5%) in the dantrolene group received intensive care unit (ICU) level care following the procedure compared to two participants (7.7%) in the placebo group. Of note, three of the five participants who required ICU-level care in the dantrolene group had already been receiving that level of care prior to the ablation procedure and the two patients who were transferred to the ICU were assigned dantrolene but were withdrawn from the trial prior to initiation of the study protocol due to complex, lengthy procedure in one case and concern for cardiogenic shock in the other. No drug-related SAEs were noted. Procedural complication rates and potential drug-related side effects were low without apparent difference between groups **(Supplemental Table 5).** No participants died during the index hospitalization in either group.

### Dantrolene pharmacokinetics

Dantrolene blood concentrations of twenty-nine participants were included in the pharmacokinetic analysis. Peak dantrolene concentrations immediately following the intravenous bolus were 4067 ± 2107 ng/ml (arterial) and 2645 ± 831ng/ml (venous). Arterial and venous concentrations equilibrated with each other within ten minutes. Twenty minutes after drug administration, mean arterial and venous dantrolene concentrations were 1477 ± 355 and 1435 ± 323 ng/ml, respectively. Following a rapid distribution phase, dantrolene exhibited a mean elimination of 17.1 hours in this population.

## DISCUSSION

Despite the urgent need for new AADs, none have been approved for over 20 years. Furthermore, among the current AADs, most are contraindicated in patients with structural heart disease and other comorbidities such as renal dysfunction, QT prolongation and conduction disease. Our results suggest that RyR2 inhibitors such as dantrolene can be considered a new class of AADs that can prevent VT in a high-risk population with significant structural heart disease and that may not have adverse electrophysiological and hemodynamic side effects.

### RyR2 Inhibition Decreased VT Inducibility

Dantrolene reduced inducible VT by 77%, whereas there was no change in VT inducibility following placebo administration (Figure 1). Although our sample size is relatively small, the double blinded, randomized design is a strength with statistically robust results. There are several relevant caveats. Our patients were at high risk for ventricular arrhythmias, but were studied after ablation was performed. Inducible arrhythmias following ablation are common, because patients with scar related VT often have multiple potential reentry circuits, not all of which may be ablated. Although these arrhythmias may not be considered “clinical”, they do reflect presence of an arrhythmia substrate and are associated with an increased chance of spontaneous sustained VT during follow-up.^19,20^

The rapid onset of action and tissue equilibrium of dantrolene in myocardial cells is consistent with its high lipophilic properties and the high rate of blood flow to the heart. The elimination half-life of dantrolene in the VT population (∼17 hours) was longer than that reported in healthy volunteers (∼11 hours). Importantly, dantrolene venous concentrations during the EP procedure (1435 – 2645 ng/ml) were 4.7 to 8.5-fold lower than concentrations required to treat malignant hyperthermia (6700–22600 ng/ml). Collectively these findings indicate that lower intravenous doses of dantrolene are sufficient for cardiac RyR2 target engagement and will likely provide a greater safety margin to treat cardiac arrhythmias.

Our results extend to humans those observed with dantrolene in preclinical models. Zamiri *et al.* used burst ventricular pacing to induce VF in 26 pigs with structurally normal hearts that were randomized to treatment with dantrolene or placebo during cardiopulmonary resuscitation.^14^ Dantrolene facilitated successful defibrillation such that 85% of pigs in the dantrolene group had restoration of sinus rhythm, compared to only 54% in the placebo group. In mice with chronic post-ischemic heart failure 4 weeks after left coronary ligation,^21^ VT was inducible in 53% of mice treated with placebo compared to only 20% of mice treated with dantrolene. The VT prevention with dantrolene observed in our randomized clinical trial is also consistent with a recent report of an uncontrolled case series of 10 patients treated with dantrolene to reduce VT storm.^16^

### Mechanism of antiarrhythmic action by dantrolene

Intracellular calcium leak from the sarcoplasmic reticulum via RyR2 channels has been widely documented in animal models and humans with heart failure and ischemia causes, which can be prevented by treatment with dantrolene.^22^ The arrhythmia mechanism most strongly linked to calcium leak are delayed afterdepolarizations (DADs).^23^ DADs can trigger ventricular ectopy and can also alter local tissue conduction, which promotes reentrant arrhythmias.^24,25^ While we cannot exclude that triggered activity contributed to induced arrhythmias, scar-related reentry is the usual cause of inducible VT associated with structural heart disease.^26,27^ Our findings suggest that dantrolene can prevent reentrant VT. AADs are thought to prevent reentrant arrhythmias either by prolonging refractoriness or suppressing conduction to the point of block in the reentry circuit.^7^ To investigate these possibilities, we assessed refractoriness and conduction in low voltage areas where myocytes and fibrosis coexist, and in higher voltage areas where less fibrosis is presumably present. Interestingly, we did not find evidence for a change in these electrophysiologic parameters in either region. We also did not detect a difference in PR interval, QRS duration or QT interval, consistent with the absence of an effect at these slow rates. These results suggest that dantrolene acts locally on small myocardial tissue bundles that contribute to reentry circuits without affecting global electrophysiological properties of the heart. Hence, Dantrolene is different from traditional class I or class II AADs, which can be pro-arrhythmic due to altering conduction or refractoriness globally in the heart, making dantrolene likely a safer AAD choice in patients with structural heart disease.

### Safety: No Drug Related SAEs or Change in ECG Intervals or Hemodynamics

There were no drug-related significant adverse events observed in this study. The lack of effect on heart rate, PR, QRS, and QT intervals would be consistent with a low risk of pro-arrhythmia from bradycardias, torsade de pointes, or VT. A favorable safety profile with no changes in ECG intervals was also reported in a study of 20 patients treated for over 10 years with chronic oral dantrolene for muscle spasticity disorders.^28^ Due to its inhibition of *skeletal muscle* RyR1 channels, Dantrolene reduces skeletal muscle strength.^29^ Theoretically, inhibition of *cardiac* RyR2 could have a negative inotropic effect that would be particularly concerning in patients with structural heart disease. Although the median LVEF in our study was reduced (35%), we did not observe significant changes in blood pressure, cardiac index, right atrial pressure, or PA pressures with administration of dantrolene. The lack of hemodynamic deterioration with dantrolene reported here is consistent with a case series finding normal LV function after prolonged dantrolene treatment in 10 patients,^28^ and with results from experimental heart failure models.^22,30,31^ Furthermore, dantrolene significantly improved hemodynamic recovery when given to pigs during cardiac arrest, and in mice with chronic ischemic heart disease.^14,21^ Our data support the hemodynamic safety of dantrolene for patients with LV systolic dysfunction.

### Limitations and future directions

Our study population was at a high risk for VT.^32^ The cardiac milieu of patients undergoing ventricular arrhythmia ablation is complex. Approximately a third of our subjects were intubated, 47% were on amiodarone, 78% were on beta blockers, and 98% were on vasopressors, which are all potential confounders for measurement of arrhythmia inducibility, hemodynamics, ECG intervals, VERP, and conduction metrics. Nevertheless, the significant difference in VT inducibility that was detected between dantrolene and placebo despite these potential confounders argues for the robustness of dantrolene’s effect at reducing VT inducibility. Prior to starting the trial, the planned enrollment was 84 participants to be completed by June 2023. The first participant was enrolled in December 2020 and then recruitment was halted and later remained slowed by the COVID-19 pandemic. To partially address this, the DSMB recommended to expand eligibility to include participants with structural heart disease undergoing catheter-ablation for premature ventricular contractions (PVCs) in April of 2021, and in August 2023 to discontinue measurement of conduction velocity to shorten case duration which would allow enrollment of patients scheduled for later in the day (who were otherwise excluded to attempt to meet restrictions on laboratory end times). Furthermore, one year of recruitment was added. In consultation with the DSMB, the trial was stopped in March 2024. Fortunately, the study was powered sufficiently to detect a significant association in the primary endpoint of inducibility, but the smaller sample size may have affected the ability to detect a significant association in secondary endpoints and to perform multivariable analyses.

Our results support further investigation into the therapeutic potential of dantrolene and the development of RyR2 specific inhibitors. Specifically, dantrolene (Ryanodex) was administered as a small volume (4 mL) I.V. bolus with a rapid time of onset (∼0.5-5 minutes) and was found to decrease VT inducibility in patients already treated with amiodarone and other AADs. It had a favorable safety profile in patients with structural heart disease and numerous comorbidities. Taken together, RyR2 stabilization holds potential as a new antiarrhythmic drug approach that could be an adjuvant medication for ACLS (advanced cardiac life support) or for patients in VT storm.

An oral formulation of dantrolene exists that has been administered chronically. Concern exists, however, over dantrolene’s RyR1 inhibition that may result in muscle weakness, respiratory depression, and hepatotoxicity. The effective dose of dantrolene for treatment of arrhythmias is not known. Interestingly, we used a lower dose in our study, 1 mg/kg, than the maximally approved dose for malignant hyperthermia, which is 2.5mg/kg. We found 1 mg/kg successfully suppressed VT inducibility, which suggests the possibility that lower doses of dantrolene could be used for treatment of arrhythmias and potentially minimize the side effects associated with RyR1 inhibition at higher doses. A randomized controlled trial has been proposed to study oral dantrolene for prevention of cardiovascular death, heart failure hospitalization, and VT/VF.^33^ Investigational RyR2 specific inhibitors that have shown promising results in pre-clinical studies and are not expected to have skeletal muscle effects could be considered for further therapeutic development.^34–36^

## METHODS

### Enrollment

We conducted a randomized, double-blinded, placebo-controlled trial from December 2020 to March 2024. Eligible participants had structural heart disease and were scheduled for a catheter-based ablation of VT or premature ventricular contractions (PVC) at Vanderbilt University Medical Center (tertiary, academic medical center). Patients had to be at least 18 years of age, able to give written informed consent, and have an implantable cardiac device capable of providing back-up pacing (pacemaker or transvenous defibrillator). Major exclusion criteria were mechanical ventricular support, New York Heart Association Class IV heart failure, left ventricular ejection fraction (LVEF) <20%, glomerular filtration rate (GFR) <30 mL/min/1.73 m^2^, body mass index (BMI) ≥40 kg/m^2^, chronic liver disease (Child Pugh Class A-C), current use of a calcium channel blocker, a neuromuscular disorder (e.g., muscular dystrophy), or chronic lung disease requiring oxygen therapy or a history of intubation. See **Supplemental Table 1** for the full set of eligibility criteria.

### Electrophysiologic Study and Ablation

VT/PVC ablation was performed according to standard clinical practice (**Supplemental Figure 1**). Oral arrhythmic medications were held for 24-48 hours. Continuous intravenous lidocaine and amiodarone were held for 4-6 hours. The PVC/VT ablation procedure was performed under monitored anesthesia care (MAC) or general anesthesia. The left ventricle was accessed via either retrograde aortic or trans-septal access. Hemodynamics were monitored with a radial or femoral arterial line and a pulmonary artery catheter. Standard electrophysiology catheters were used: 1) a quadripolar catheter placed in the right ventricle (RV); 2) a multipolar mapping catheter (Decanav, Biosense Webster Inc., Irvine, CA); and 3) an irrigated-tip radiofrequency ablation catheter (Thermacool SmartTouch SF or QDOT Micro Catheter, Biosense Webster Inc., Irvine, CA). Cardiac mapping was performed using CARTO-III (Biosense Webster Inc., Irvine, CA). Intracardiac echocardiography (ICE) was utilized in all patients for defining anatomy, assessing catheter position and observing for complications.

### Randomized Study of Dantrolene

For each enrolled participant, the study protocol was started after completion of the clinical ablation procedure. Participants were randomized 2:1 to dantrolene (treatment arm) or placebo (control arm). We used a stratified randomly-permuted block randomization, with block sizes varying randomly from 4 to 6 to ensure overall balance between treatment arms. Randomization was stratified by 1) amiodarone use within the preceding 21 days; 2) LVEF <35%; and 3) PVC vs. VT ablation. The participants and members of the study team performing electrophysiologic and hemodynamic measurements were blinded to the treatment group. Personnel who enrolled and assigned participants to the treatment group did not have access to the random allocation sequence.

We quantified conduction velocity and the effective refractory period in two regions: Areas where the bipolar electrogram amplitude was < 1.5 mV (low voltage), which are areas that are a combination of myocytes and fibrosis, and areas of bipolar voltage > 1.5 mV (higher voltage) that have less or possibly no fibrosis **(Supplemental Figure 2)**. Pacing at twice diastolic threshold at a basic cycle length of 600 ms for 8 beats was followed by a single extrastimulus followed by a 4 second pause. The effective refractory period (ERP) was defined as the coupling interval with loss of capture on two consecutive attempts. Conduction times were measured offline by a blinded reviewer and defined as the time from the stimulus artifact to the dominant peak of the bipolar electrogram on the electrode most distant from pacing site of pacing that had a consistent, reliable electrogram. When multiple, small signals were present, as often found near scar, the latest sharp peak was used. Conduction restitution curves were created by plotting conduction time versus coupling interval of the premature stimulus and used to determine the average basal conduction time and the inflection point defined as the longest coupling interval at which the conduction time exceeded that of the baseline conduction time by 5 ms and continued to increase **(Supplemental Figure 3)**.^17^

VT inducibility was determined utilizing a standardized VT protocol, progressing from 1 to 3 extrastimuli following a drive train of 600 ms from the RV apex (**Supplemental Methods 1**).^18^

Hemodynamic measurements included mean arterial pressure (MAP), right atrial (RA) pressure, pulmonary artery (PA) systolic pressure, PA diastolic pressure, and cardiac index (CI). CI was calculated by the Fick method using PA mixed venous and arterial oxygen saturations. The research protocol was stopped if the pre-study drug PA oxygen saturation was <50%.

Respiratory mechanics were continuously monitored by the anesthesiology team and a neuromuscular and respiratory mechanics sub-study will be reported separately.

Participants received the study drug (dantrolene or placebo) administered intravenously in 4 divided boluses over 3 minutes. An ECG was continuously recorded, and hemodynamic measurements were obtained every 5 minutes. The average of 3 consecutive PR, QRS, RR and QT intervals were measured off-line every 2 minutes by a reviewer blinded to treatment assignment. Twenty minutes after study drug administration, measurements of conduction and refractoriness were repeated in both the low and higher voltage regions followed by the VT induction protocol.

### Study Drug and Pharmacokinetic Analysis

Dantrolene was formulated as a nanoparticle suspension (RYANODEX, Eagle Pharmaceuticals Inc., Woodcliff Lake, NJ). Compared to conventional formulations of 1dantrolene, Ryanodex has enhanced aqueous solubility that allows rapid intravenous injection and is approved for treatment of malignant hyperthermia at doses ranging from 1 mg/kg to 2.5 mg/kg. Peak concentration is observed 1 minute after drug administration and the mean half-life in healthy individuals is 10 hours. Based on its safety and measurable pharmacodynamic effect (i.e., reduced grip strength) in an open label pilot study (N=5 participants), we chose the lowest dose, 1 mg/kg, for our study.

To evaluate the pharmacokinetic properties of dantrolene in this study population, femoral venous and arterial blood samples were collected at baseline and at 5, 10, 15, and 20 minute-intervals post-drug administration. Four additional blood samples were randomly collected post-ablation at defined intervals (1-4 hours, 6-12 hours, 16-24 hours, 28-48 hours). Plasma samples were stored at −80°C. Dantrolene concentrations were measured by liquid chromatography-tandem mass spectrometry using multiple reaction monitoring.

### Efficacy endpoint

The primary efficacy outcome was VT/VF inducibility, which was defined as sustained VT/VF (>= 10 seconds), VT/VF requiring termination (with pacing or defibrillation), or two or more episodes of non-sustained VT lasting 3-9 seconds. It was analyzed as a binary variable (yes/no) and an ordinal measure (1-6) based on the stage of the induction protocol at which the primary endpoint was met.

### Safety endpoints

The safety endpoints were hemodynamics, myocardial electrophysiology and other adverse drug events. Hemodynamic safety parameters included mean arterial pressure (MAP), post-drug cardiac index, and PA systolic and diastolic pressures. Electrophysiological safety parameters included post-drug conduction time and ERP in low and higher voltage areas, and ECG intervals. The safety analysis set included all randomized participants who received dantrolene. Serious adverse events (SAEs) were defined as those that resulted in death, were life-threatening, required inpatient hospitalization or prolonged hospitalization, and/or resulted in permanent or significant disability. We defined a drug-related adverse effect as one that occurred during or after administration of the study drug with at least one of the following criteria: 1) the event could not be explained by the clinical condition or history, environmental factors or other diagnostic and therapeutic measures; 2) was an expected adverse effect associated with the study drug or a class-labeled drug effect; or 3) the adverse effect subsided or disappeared after withdrawal of the study drug.

### Statistical Analysis

Baseline characteristics are summarized descriptively by treatment group. Categorical data are presented as frequencies and percentages and analyzed with Chi-squared test. Continuous data are summarized with mean (SD) and median (Q1, Q3). To address the number of participants who were withdrawn due to case duration and complexity during the clinical VT ablation, but prior to study drug administration or the research protocol, the DSMB requested in August 2023 changing the analysis to a Modified “Intention-to-Treat (mITT). The mITT analysis was performed such that all randomized participants who underwent VT/PVC ablation and received any study treatment (dantrolene or placebo) were analyzed according to the study arm they were originally assigned. Between-group differences were assessed using Wilcoxon rank sum test or Kruskal-Wallis test. Continuous outcomes with repeated measurements (e.g., heart rate, blood pressure, arterial pressure, cardiac index, and corrected QT interval) were analyzed with linear mixed-effect models to account for potential correlation in the data. Less frequent post-treatment measurements such as RA and PA pressures were analyzed using linear regression models adjusted for pre-treatment levels. The primary outcome of the trial was VT inducibility. VT inducibility was analyzed as a binary variable (yes/no) and ordinal variable. Univariable associations were explored between VT inducibility with amiodarone exposure (yes/no), estimated GFR, pre-procedure LVEF, BMI, history of prior myocardial infarction and norepinephrine dose. To correct for multiple comparison testing, the P-value threshold for statistical significance was adjusted using Bonferroni’s, which applied to the analysis of the baseline demographics, clinical characteristics, and secondary outcome measurements. No imputation was performed for missing data. All statistical analyses were performed in R version 4.4.3.

The power calculation indicated that 84 participants were required to achieve 80% power to detect an odds ratio (OR) of 0.25 for the primary endpoint of inducibility (yes/no) at a two-sided significance level of 5%.

De-identified summary measures such as overall baseline characteristics, as well as outcomes summary will be made openly available. De-identified individual level data will be available through a controlled access plan requiring a confidentiality agreement and institutional approval. The trial protocol, data dictionary, and analysis plan can be made available upon request to the corresponding author.

## ACKNOWLEDGEMENTS

We thank the Vanderbilt University Medical Center Electrophysiology Laboratory staff for their professionalism and dedication, which makes research like this possible.

## AUTHOR CONTRIBUTIONS

MAE acquired, analyzed, and interpreted data, and drafted and revised the manuscript. ZTY assisted with conception and design of the study. BV performed data acquisition and analysis. DG, DMC, and HL assisted with data acquisition, data management, and drug randomization/administration. FY and LS assisted with study design and performed statistical analysis and data interpretation. JS, HT, TR, AK, GD assisted with data acquisition and interpretation. DR assisted with study design and provided critical feedback on the manuscript. MM, MP, FTB, KS performed data acquisition and analysis for the neuromuscular and respiratory portion of the study. WSA performed data analysis for the pharmacokinetic/dynamic portion of the study. BCK, WGS, and MBS conceived and designed the study, performed data acquisition, analysis, and interpretation, and critically revised the manuscript. All authors reviewed and provided feedback on the manuscript.

