## Supplementary material for "Ryanodine Receptor Inhibition with Dantrolene Prevents Ventricular Tachycardia Induction in Patients with Structural Heart Disease – A Randomized Controlled Trial": Data Supplement

**SUPPLEMENTAL TABLES AND FIGURES**

**Supplemental Table 1: Eligibility Criteria**

**Supplemental Figure 1: Study Protocol**

**Supplemental Figure 2: Electroanatomic Bipolar Voltage Map of the Left Ventricle**

**Supplemental Figure 3: A Conduction Restitution Curve.**

**Supplemental Methods 1: Induction Protocol**

**Supplemental Figure 4: Consort Diagram**

**Supplemental Table 2: : Baseline characteristics and ablation of all randomized participants**

**Supplemental Figure 5: Mean arterial blood pressure before and after study drug**

**Supplemental Figure 6: Cardiac index before and after study drug**

**Supplemental Figure 7: Mean arterial blood pressure over time**

**Supplemental Figure 8: Change in cardiac index**

**Supplemental Figure 9: Change in RR interval**

**Supplemental Figure 10: Change in PR interval**

**Supplemental Figure 11: Change in QRS duration**

**Supplemental Figure 12: Change in corrected QT interval**

**Supplemental Figure 13: QRS duration before and after study drug**

**Supplemental Figure 14: QTc duration before and after study drug**

**Supplemental Table 4: Ventricular Repolarization and Conduction**

**Supplemental Table 5: Safety Outcomes and Adverse Events**

**Supplemental Table 1: Eligibility Criteria**

**Inclusion**

≥18 years of age

Able to give written, informed consent

Referred for catheter-based VT ablation or PVC ablation

Structural heart disease (LVEF < 50%, history of prior myocardial infarction, cardiac sarcoid or valvular heart disease)

Permanent pacemaker or implantable cardioverter defibrillator

**Exclusion**

Mechanical ventricular support (e.g. LVAD, ECMO)

NYHA class IV heart failure

LVEF <20%

Severe renal insufficiency (GFR<30 mL/min)

Morbid obesity (BMI≥ 40 kg/m^2^)

Chronic liver disease (Child Pugh class A-C)

Current use of calcium channel blockers

Neuromuscular disorder (e.g. muscular dystrophy)

Chronic obstructive pulmonary disease or restrictive lung disease requiring oxygen therapy or history of intubation

Pregnant or nursing

History of dysphagia

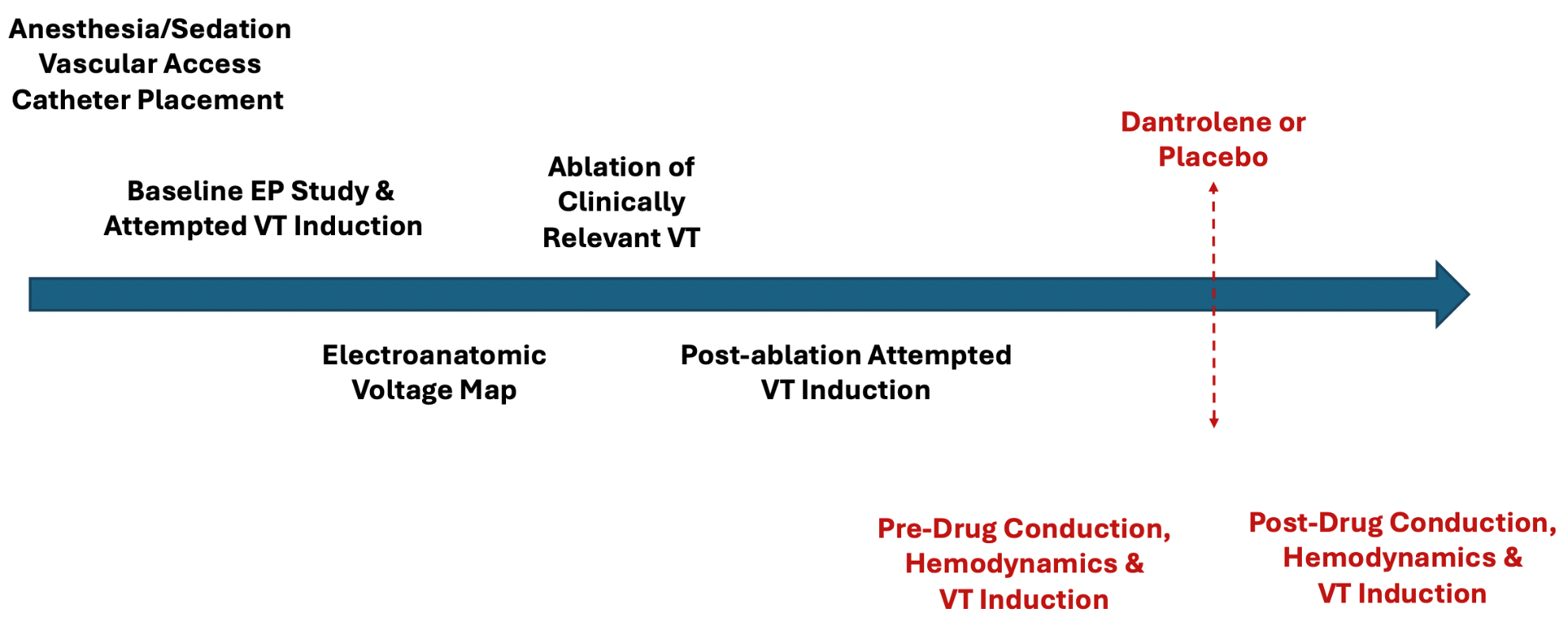
**Supplemental Figure 1: Study Protocol.** The study protocol (red) was integrated into the clinical workflow of catheter-based ventricular arrhythmia ablation. Shown here is the workflow for VT ablation.

**Supplemental Figure 2: Electroanatomic Bipolar Voltage Map of the Left Ventricle:** The participant has an anterior wall infarct. Colors from red, yellow, green, blue and purple indicate progressively higher voltage. Red (**A**) is dense scar less than 0.1 mV. The scar border zone is near the edge where cardiomyocytes are mixed with fibrosis (**B**). Conduction and ventricular effective refractory periods were measured by multipolar catheters in the scar border zone (**C**) and higher voltage tissue (**D**).

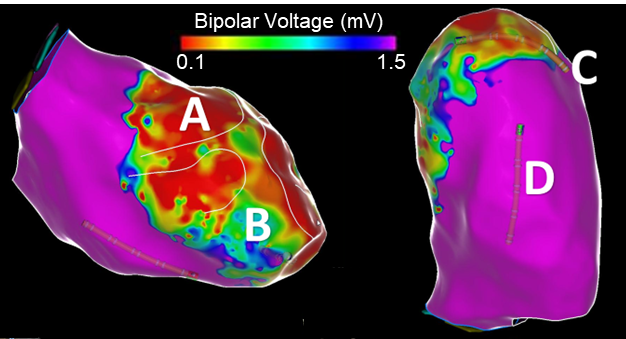

**Supplemental Figure 3: A Conduction Restitution Curve:** The conduction time is measured from pacing at a proximal electrode to the local electrogram recorded at a distal electrode and plotted against the coupling interval (S2). **Label A.** The basal conduction time. **Label B**. The inflection point of the conduction restitution curve.

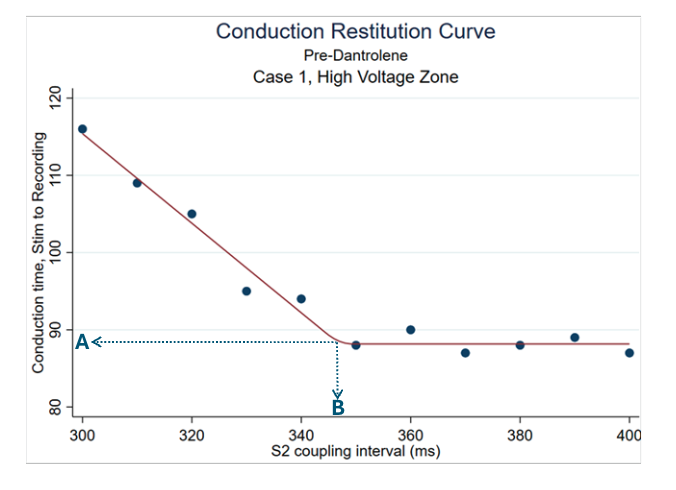

**Supplemental Methods 1: Induction Protocol**

Pacing stimulus strength was twice the diastolic threshold. The pacing cycle length (S1-S1) was 600 ms for 8 beats followed by extrastimuli (ES) and a 4 second pause. The first extrastimulus (S2) was introduced at a coupling interval 400 ms and decremented by 10 ms to VERP or 200 ms. S2 was then set to a coupling interval (S1 – S2) of VERP (or 200 ms) + 20 ms and a second extrastimulus (S3) added at a coupling interval of S1-S2 + 50 ms and scanned to refractoriness or a minimum coupling interval of 200 ms. The second extrastimulus was then set to its VERP + 20 ms and a third extrastimulus (S4) added at an initial coupling interval (S3 – S4) of S2 – S3 + 50 ms and scanned to refractoriness or 200 ms. Thus, up to 3 extrasimuli were added. The minimum coupling interval was no shorter than 200 ms. The induction protocol stopped when: 1) the primary endpoint was met, or 2) the induction protocol had been completed. The primary endpoint was an episode of sustained VT that lasted ≥10 seconds or two episodes of non-sustained ventricular arrhythmia defined as VT or VF that lasted 3-9 seconds.

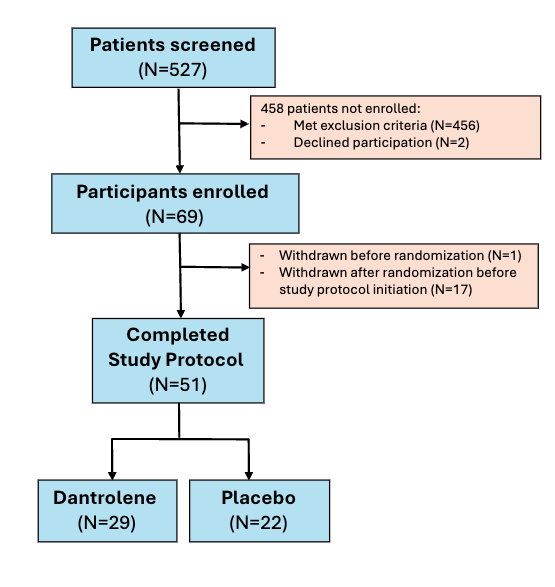
**Supplemental Figure 4: Consort Diagram**

**Supplemental Table 2: :** Baseline characteristics and ablation of all randomized participants

|  | **Total (N=68)** | **Dantrolene (N=41)** | **Placebo (N=27)** | **P-value** |
| --- | --- | --- | --- | --- |
| Age at enrollment  Male Sex  Race  White  Black  Asian  Ethnicity- Non-Hispanic  Height (cm)  Weight (kg)  Body mass index (kg/m2)  Hypertension  Atrial Fibrillation  COPD  Coronary Artery Disease  Chronic Kidney Disease  Amiodarone Use within 21 days  Pre-procedure Creatinine (mg/dl)  Pre-procedure Ejection Fraction (%)  Estimated Glomerular Filtration Rate  Ablation Indication  VT  PVC  Oral antiarrhythmic within 3 days  Amiodarone  Sotalol  Dronedarone  Dofetilide  Class 1C Agents  Quinidine  Mexiletine  Intravenous antiarrhythmic within 3 days  Amiodarone  Lidocaine  Procainamide  Oral beta-blocker within 3 days  Epicardial Access  Procedure duration (hours)  Time to research protocol  Time to drug administration  Endotracheal Intubation  Pre-Study Drug Vasopressor Use  Norepineprhine  Epinephrine  Vasopressin  Phenylephrine  Mean Norepinephrine Dose (mg/min) | 68.4 (61.6-74.3)  63 (92.6%)  60 (88.2%)  7 (10.3%)  1 (1.5%)  68 (100.0%)  180.3 (175.3-183.5)  99.0 (80.6-109.1)  30.0 (26.2-32.7)  57 (83.8%)  27 (39.7%)  11 (16.4%)  41 (60.3%)  12 (17.6%)  32 (47.1%)  1.0 (0.9-1.2)  35.5 (30.0-45.0)  76.6 (61.2-91.8)  45 (88.2%)  6 (11.8%)  31 (60.8%)  18 (35.3%)  9 (17.7%)  0  0  0  1 (2.0%)  14 (27.5%)  10 (19.6%)  1 (2.0%)  11 (21.6%)  0  40 (78.4%)  9 (17.6%)  5.8 (4.4-7.1)  3.6 (2.5-4.6)  4.0 (3.1-5.1)  17 (33.3%)  50 (98.0%)  42 (82.4%)  4 (7.8%)  4 (7.8%)  8 (15.7%)  5.0 (3.2-6.8) | 69.0 (61.7-74.4)  38 (92.7%)  36 (87.8%)  4 (9.8%)  1 (2.4%)  41 (100.0%)  180.3 (177.8-185.4)  98.9 (90.7-109.0)  30.8 (27.7-33.1)  35 (85.4%)  17 (41.5%)  7 (17.1%)  25 (61.0%)  8 (19.5%)  20 (48.8%)  1.1 (0.9-1.3)  40.0 (30.0-45.0)  72.7 (59.8-89.3)  24 (82.8%)  5 (17.2%)  19 (65.5%)  12 (41.4%)  2 (6.9%)  0  0  0  1 (3.5%)  10 (34.5%)  6 (20.7%)  1 (3.5%)  7 (24.1%)  0  25 (86.2%)  2 (6.9%)  5.3 (4.5-6.5)  3.4 (2.6-4.5)  4.0 (3.0-5.0)  6 (20.7%)  29 (100%)  24 (82.8%)  4 (13.8%)  3 (10.3%)  5 (17.2%)  4.5 (4.0-6.2) | 66.9 (61.1-74.1)  25 (92.6%)  24 (88.9%)  3 (11.1%)  0 (0.0%)  27 (100.0%)  180.3 (175.3-182.9)  99.7 (70.9-108.8)  27.4 (24.8-32.5)  22 (81.5%)  10 (37.0%)  4 (15.4%)  16 (59.3%)  4 (14.8%)  12 (44.4%)  1.0 (0.9-1.1)  34.0 (30.0-43.8)  87.1 (68.3-97.5)  21 (95.5%)  1 (4.5%)  12 (54.5%)  6 (27.3%)  7 (31.9%)  0  0  0  0  4 (18.2%)  4 (18.2%)  0  4 (18.2%)  0  15 (68.2%)  7 (31.8%)  6.8 (4.1-7.6)  3.6 (2.5-4.6)  4.3 (3.1-5.6)  11 (50.0%)  21 (95.5%)  18 (81.9%)  0  1 (4.6%)  3 (13.6%)  5.5 (3.0-6.8) | 0.997  0.84  0.67  ---  ---  ---  0.33  0.62  0.20  0.16  0.93  0.32  0.40  0.69  0.10  0.24  0.10  0.57  0.02  0.16  ---  ---  0.73  ---  ---  ---  ---  ---  ---  ---  0.82  ---  ---  ---  0.121  0.02  0.09  0.56  0.36  0.03  0.34  ---  ---  ---  ---  0.93 |
| *P<0.002 is the threshold for statistical significance based on Bonferroni correction for multiple comparisons (0.05/27) | | | | |

**Supplemental Table 3. Ventricular Tachycardia Inducibility by Number of Extrastimuli**

|  | **Total (N=51)** | | **Dantrolene (N=29)** | | **Placebo (N=22)** | |
| --- | --- | --- | --- | --- | --- | --- |
|  | **Pre** | **Post** | **Pre** | **Post** | **Pre** | **Post** |
| VT Inducible (yes) | 22 (43.1%) | 13 (25.5%) | 12 (41.4%) | 4 (13.8%) | 10 (45.5%) | 9 (40.9%) |
| Stage that VT Induced*  S1/S2 (Single extra beats)  S1/S2/S3 (Double extra beats)  S1/S2/S3/S4 (Triple extra beats) | 1 (4.5%)  6 (27.3%)  15 (68.2%) | 1 (7.7%)  4 (30.8%)  8 (61.5%) | 0  2 (16.7%)  10 (83.3%) | 0  1 (25.0%)  3 (75.0%) | 1 (10.0%)  4 (40.0%)  5 (50.0%) | 1 (11.1%)  3 (33.3%)  5 (55.6%) |

*Given there were zero participants in the S1/S2 category for the dantrolene group and there were only a total of 13 participants that were inducible post-study drug, ordinal regression analysis could not be performed. This is because it would produce unreliable, often non-finite, standard error estimates, and the number of events falls below the accepted ratio of events-per-parameter that is required for stable calculation of the coefficient estimates.

**Supplemental Figure 5: Mean arterial blood pressure before and after study drug.** There was no reduction in mean arterial pressure (MAP) at 20 minutes after study drug administration in the dantrolene or placebo groups (both P>0.05, Wilcoxon Sign Test).

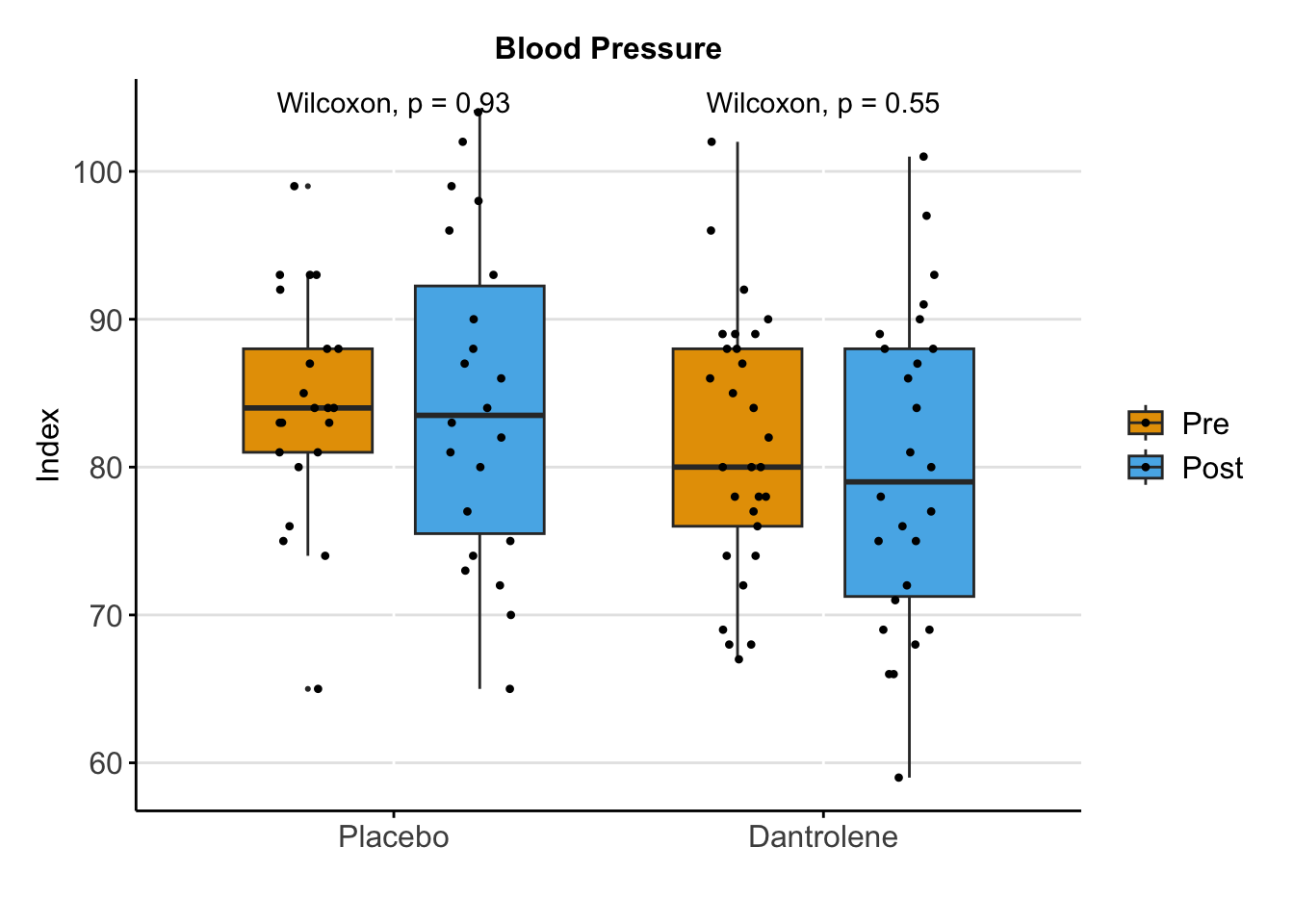

**Supplemental Figure 6: Cardiac index before and after study drug.** There was no reduction in cardiac index (Fick method) at 20 minutes after study drug administration in the dantrolene or placebo groups (both P>0.05, Wilcoxon Sign Test).

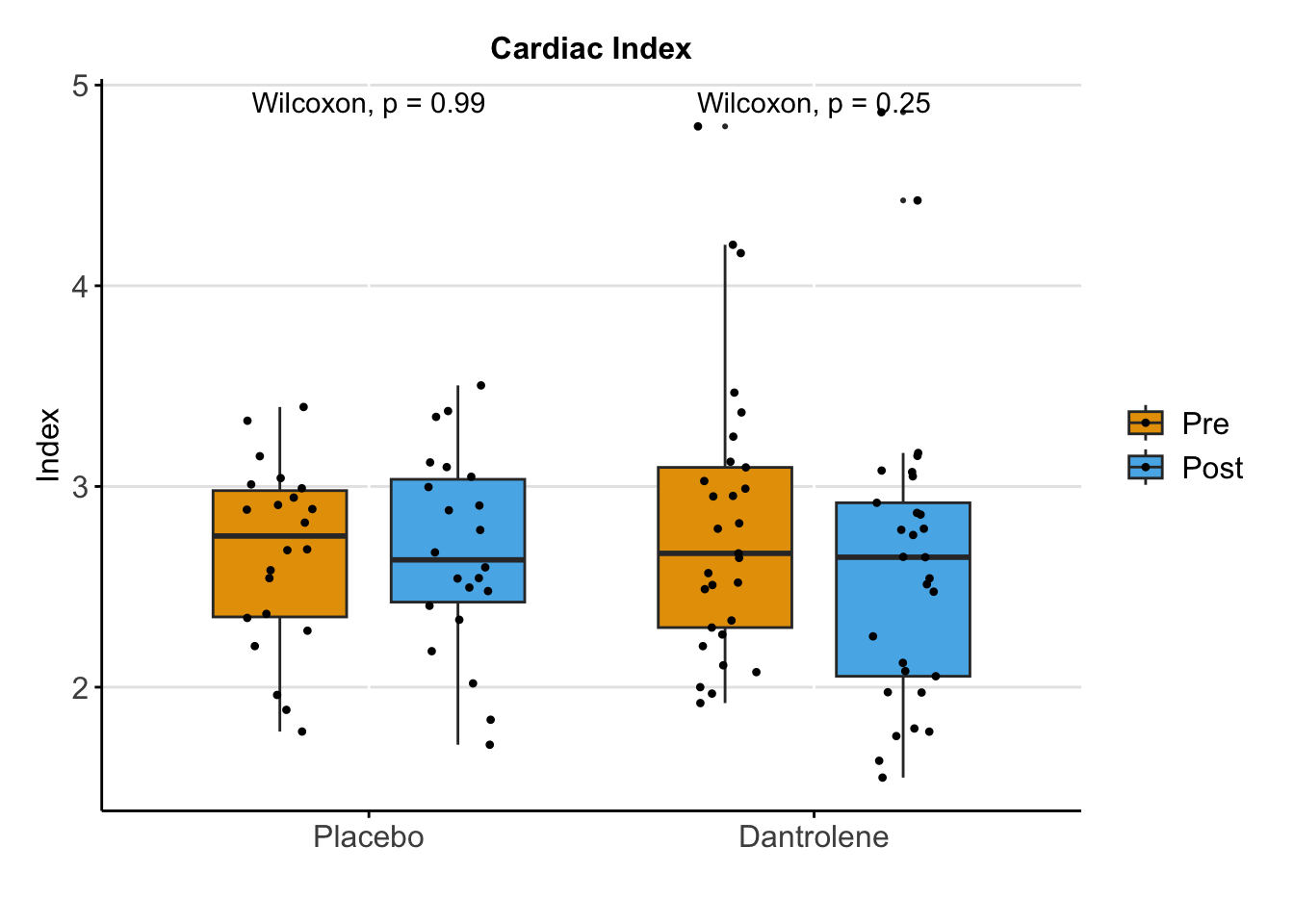

**Supplemental Figure 7: Mean arterial blood pressure over time.** There was no association between dantrolene and mean arterial pressure in a linear mixed-effect model (beta=-1.3 [95% CI -6.1, 3.6], p=0.62).

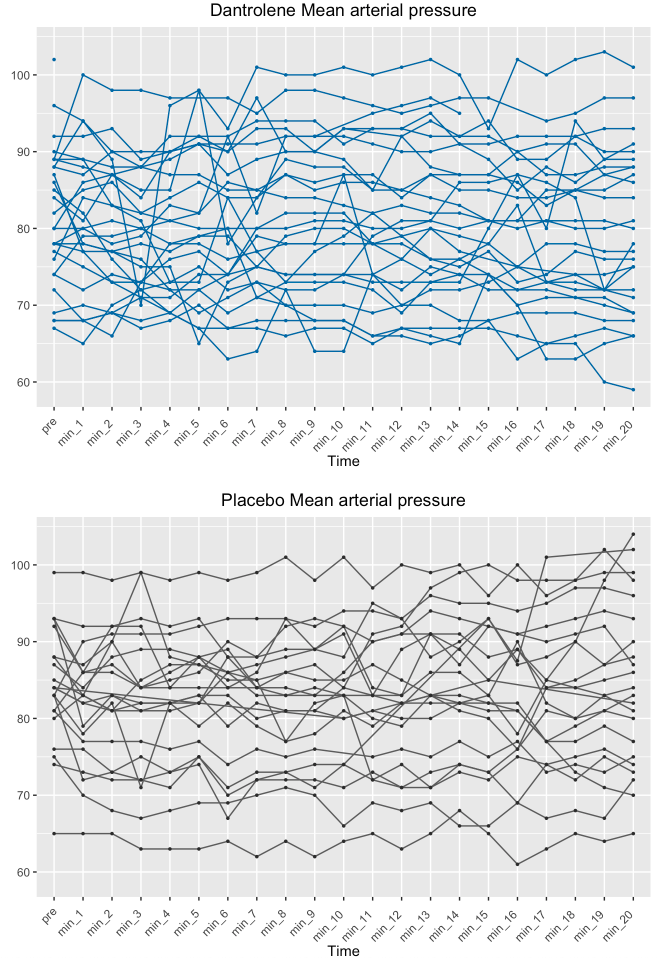

**Supplemental Figure 8: Change in cardiac index.** There was no association between dantrolene and cardiac index in a linear mixed-effect model (beta=0.2 [95% CI -0.4, 0.3], p=0.97).

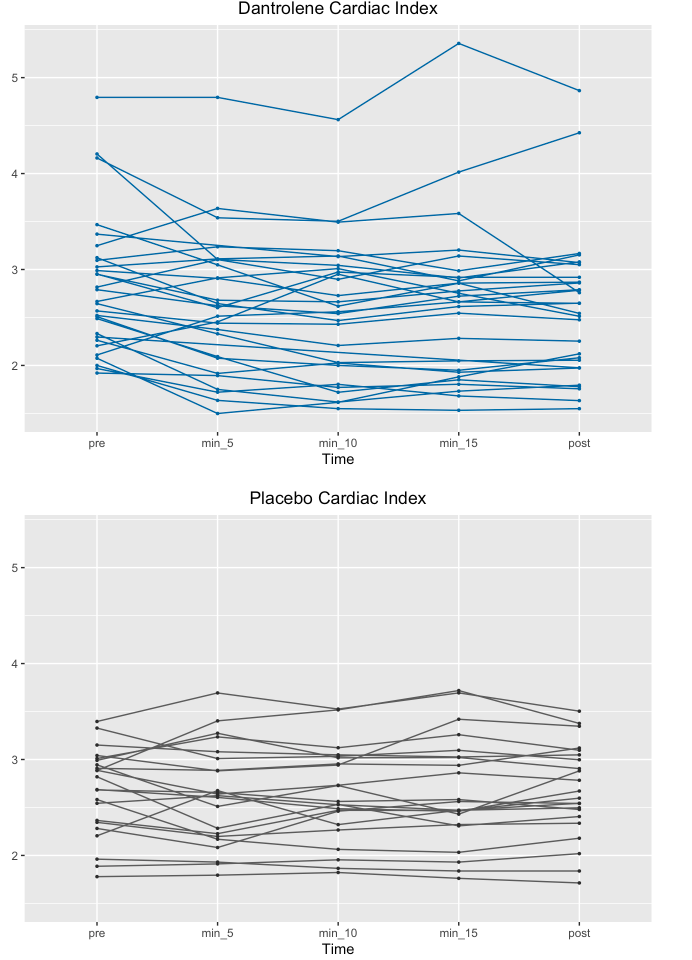

**Supplemental Figure 9: Change in RR interval.** There was no association between dantrolene and RR interval in a linear mixed-effect model (beta=-20 [95% CI -83, 43], p=0.54).

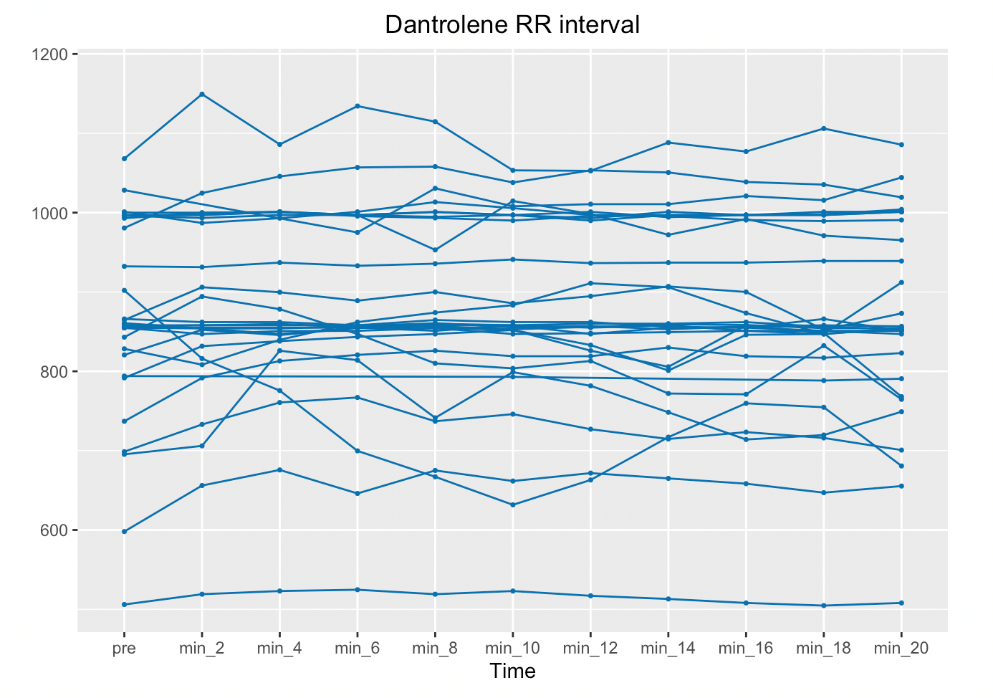

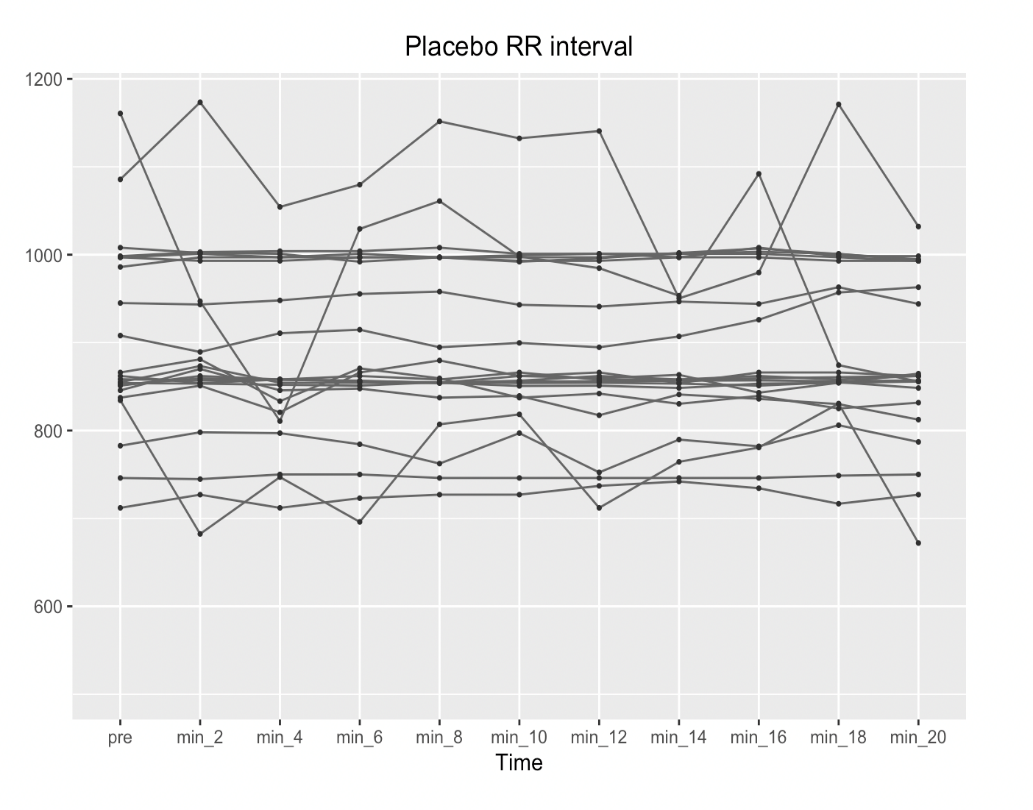

**Supplemental Figure 10: Change in PR interval.** There was no association between dantrolene and PR interval in a linear mixed-effect model (beta=2.8 [95% CI -30.4, 35.9], p=0.87).

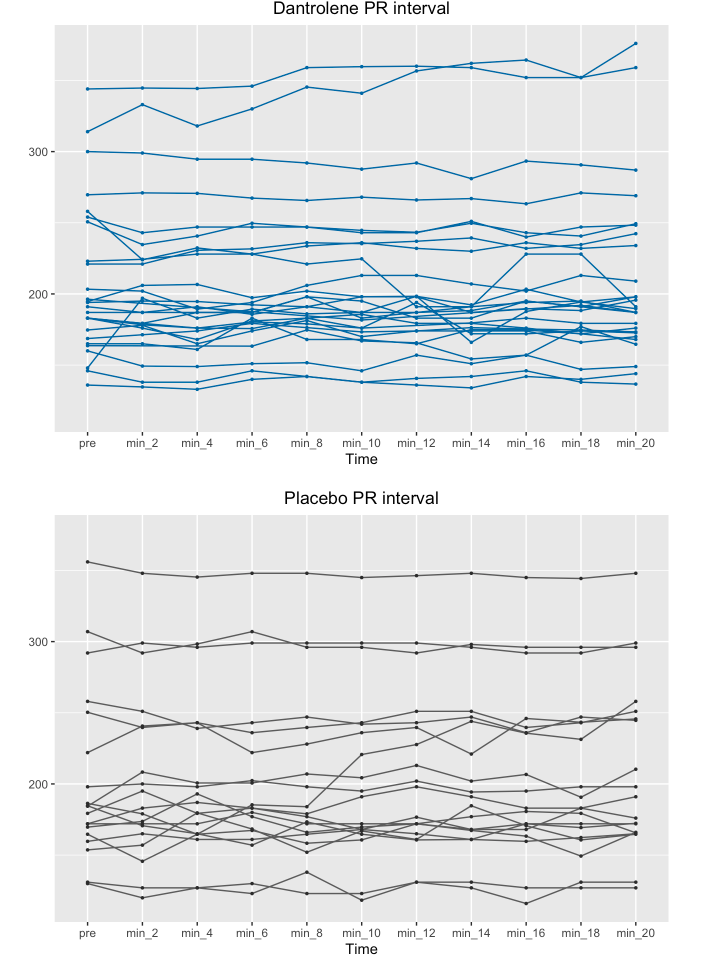

**Supplemental Figure 11: Change in QRS duration.** There was no association between dantrolene and QRS duration in a linear mixed-effect model (beta=0.4 [95% CI -19.7, 20.4], p=0.97).

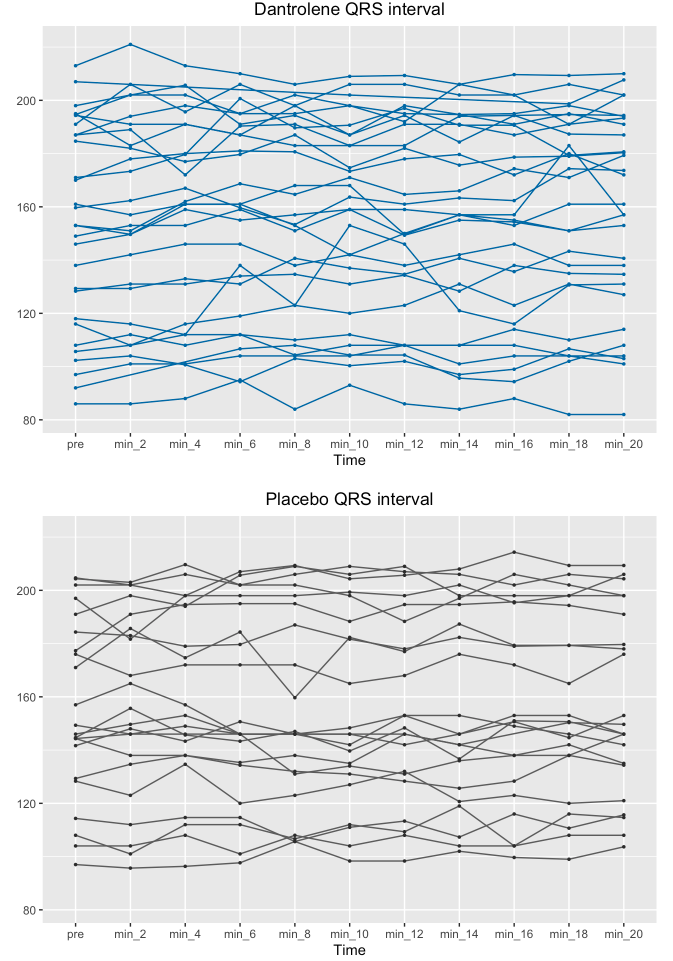

**Supplemental Figure 12: Change in corrected QT interval.** There was no association between dantrolene and QTc interval in a linear mixed-effect model (beta=-19.2 [95% CI -40.9, 2.6], p=0.09).

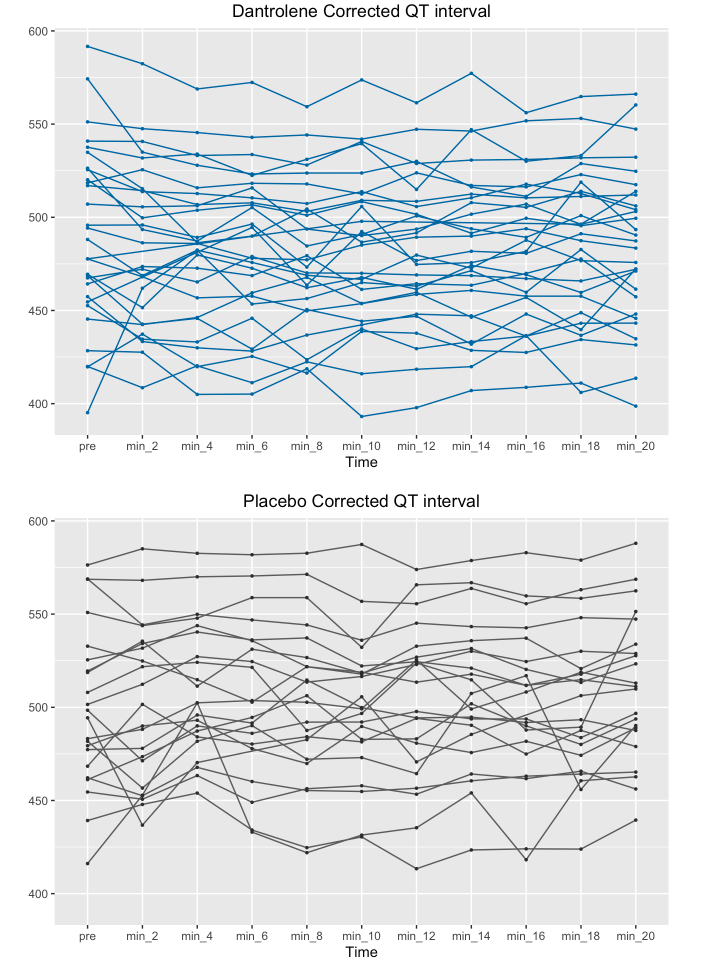

**Supplemental Figure 13: QRS duration before and after study drug.** There was no reduction in QRS duration at 20 minutes after study drug administration in the dantrolene or placebo groups (both P>0.05, Wilcoxon Sign Test).

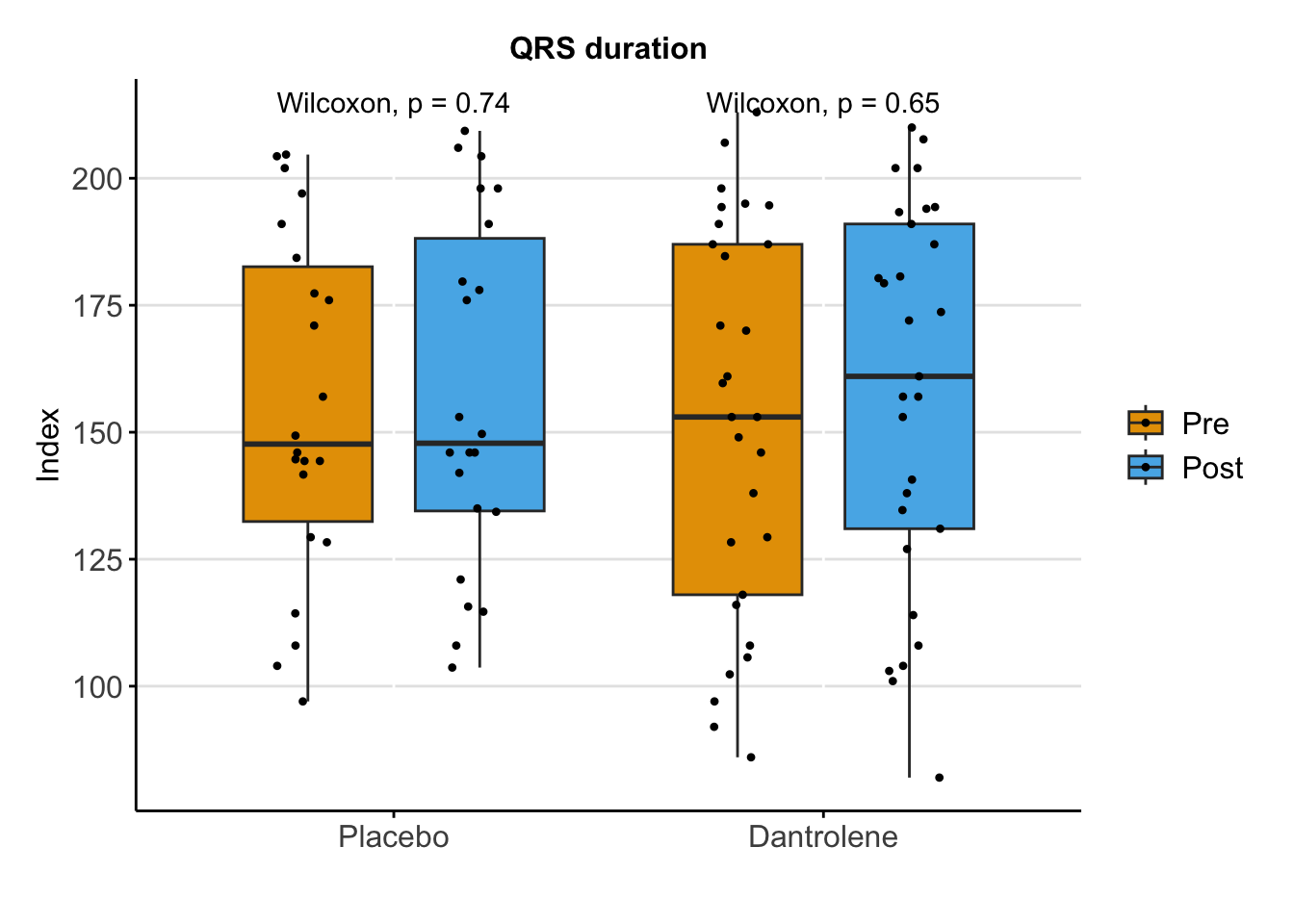

**Supplemental Figure 14: QTc duration before and after study drug.** There was no reduction in QTc interval at 20 minutes after study drug administration in the dantrolene or placebo groups (both P>0.05, Wilcoxon Sign Test).

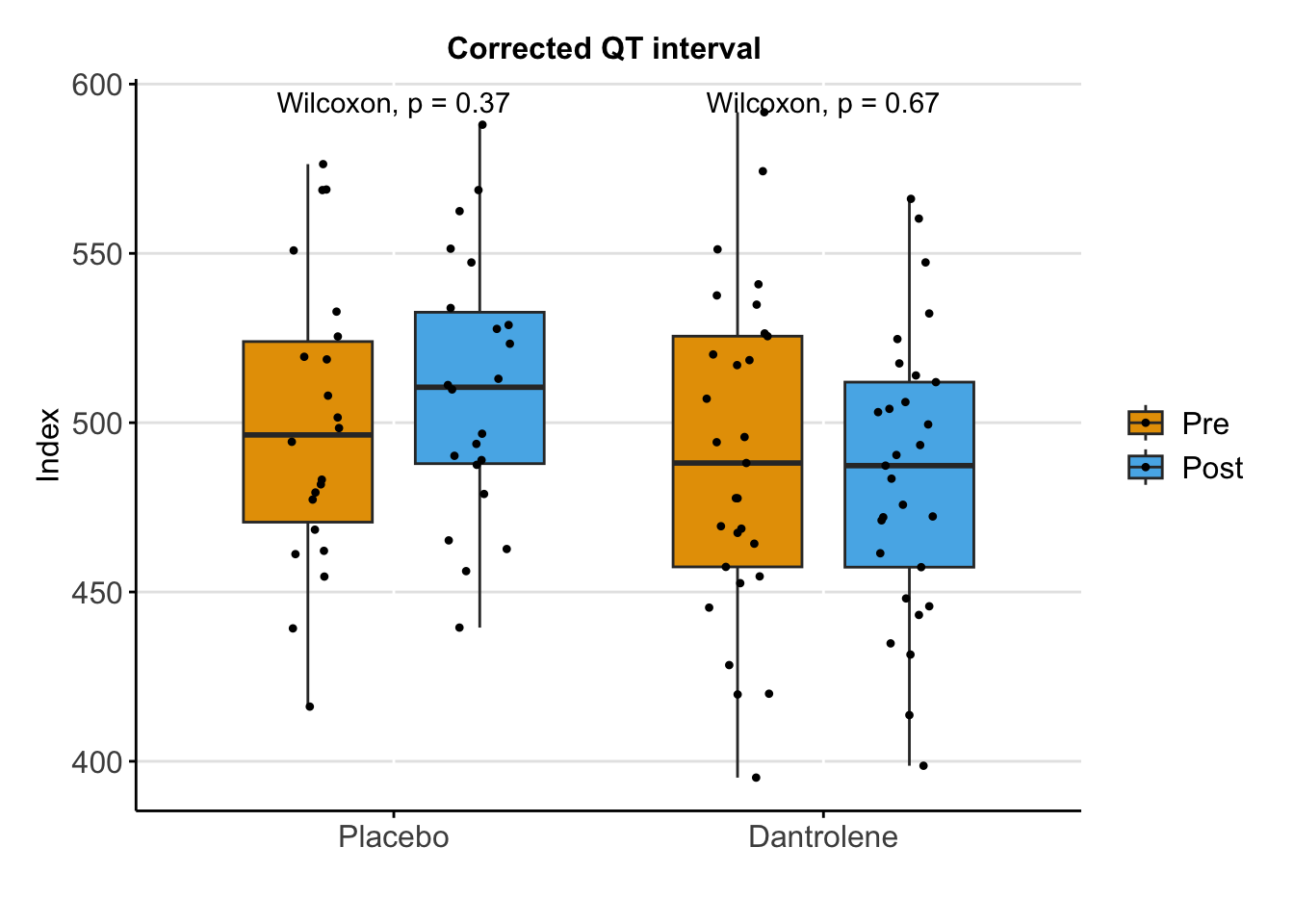

**Supplemental Table 4: Ventricular Repolarization and Conduction**

|  | **Dantrolene (N=29)** | | **Placebo (N=22)** | | **P-value*** |
| --- | --- | --- | --- | --- | --- |
|  | **Pre** | **Post** | **Pre** | **Post** |  |
| **Ventricular Effective Refractory Period (VERP)**  Higher Voltage (≥1.5 mV) Tissue  Low Voltage (<1.5 mV) Tissue | 280 (260-290)  280 (260-310) | 275 (260-290)  280 (255-340) | 280 (270-300)  290 (250-320) | 270 (260-300)  285 (260-320) | 0.99  0.91 |
| **Basal Conduction Time**  Higher Voltage (≥1.5 mV) Tissue  Low Voltage (<1.5 mV) Tissue  **Conduction Restitution Curve Inflection Point**  Higher Voltage (≥1.5 mV) Tissue  Low Voltage (<1.5 mV) Tissue | 50.8 (41.0-57.0)  66.0 (48.0-98.0)  362 (320-375)  330 (295-369) | 53.3 (44.0-69.0)  70.0 (52.0-102)  325 (305-338)  315 (301-337) | 65.0 (51.4-85.0)  87.7 (63.3-124)  344 (319-360)  340 (303-355) | 58.0 (44.3-88)  88 (62.5-110)  318 (307-333)  340 (313-354) | 0.38  0.62  0.38  0.58 |
| *****P-value for comparison of the difference in pre- vs post-study drug values (Δ post - pre) between dantrolene and placebo | | | | | |

**Supplemental Table 5: Safety Outcomes and Adverse Events**

|  | **Total (N=68)** | **Dantrolene (N=41)** | **Placebo (N=27)** |
| --- | --- | --- | --- |
| Required vasopressor support post-ablation  Intra-aortic balloon pump post-ablation  Required ICU-level care post-ablation  Procedural Complications  Hypotension  Bleeding  Cardiac Ischemia  Pericarditis  Atrial Arrhythmia  Pericardial Bleeding  Oropharyngeal bleeding  Drug Side Effects  Flushing  Somnolence  Dysphonia  Dysphagia  Nausea/vomiting  Headache  Blurred Vision  Subjective muscle weakness  Heart block  Infusion site reaction  Dizziness  Dyspnea  Urticaria  Anaphylaxis  Death within index hospitalization  Change in Norepinephrine Dose after Study Drug  Yes  No | 2 (2.9%)  1 (1.5%)  7 (10.3%)  3 (4.4%)  2 (2.9%)  1 (1.5%)  1 (1.5%)  1 (1.5%)  2 (2.9%)  1 (1.5%)  0  0  0  0  0  0  0  1 (1.5%)  0  0  1 (1.5%)  0  0  0  0  5 (9.8%)  46 (90.2%) | 1 (2.4%)  1 (2.4%)  5 (12.2%)  2 (4.9%)  1 (2.4%)  1 (2.4%)  0  1 (2.4%)  0  0  0  0  0  0  0  0  0  1 (2.4%)  0  0  1 (2.4%)  0  0  0  0  3 (10.3%)  26 (89.7%) | 1 (3.7%)  0  2 (7.4%)  1 (3.7%)  1 (3.7%)  0  1 (3.7%)  0  2 (7.4%)  1 (1.5%)  0  0  0  0  0  0  0  0  0  0  0  0  0  0  0  2 (9.1%)  20 (90.9%) |
